# Two-tiered SARS-CoV-2 seroconversion screening in the Netherlands and stability of nucleocapsid, spike protein domain 1 and neutralizing antibodies

**DOI:** 10.1101/2020.10.07.20187641

**Authors:** Anja Garritsen, Anja Scholzen, Daan W.A. van den Nieuwenhof, Anke P.F. Smits, E. Suzan Datema, Luc S. van Galen, Milou L.C.E. Kouwijzer

**Affiliations:** Innatoss Laboratories B.V., Oss, The Netherlands

**Keywords:** SARS-CoV-2, serology, lateral flow assay, ELISA, longevity, neutralizing antibodies

## Abstract

Serological testing in the COVID-19 pandemic is mainly implemented to gain sero-epidemiological data, but can also retrospectively inform about suspected SARS-CoV-2 infection. We verified and applied a two-tiered testing strategy combining a SARS-CoV-2 receptor-binding domain (RBD)-specific lateral flow assay (LFA) with a nucleocapsid protein (NCP) IgG ELISA to assess seroconversion in n=7241 individuals. The majority had experienced symptoms consistent with COVID-19, but had no access to RT-PCR testing. Longitudinal follow-up in n=97 LFA+ individuals was performed up to 20 weeks after initial infection using NCP and spike protein S1 domain (S1) IgG ELISAs and a surrogate virus neutralization test (sVNT). Individuals reporting symptoms from January 2020 onwards showed seroconversion, as did a considerable proportion of asymptomatic individuals. Seroconversion for symptomatic and asymptomatic individuals was higher in an area with a known infection cluster compared to a low incidence area. Overall, 94% of individuals with a positive IgG result by LFA were confirmed by NCP ELISA. The proportion of ELISA-confirmed LFA results declined over time, in line with contracting NCP IgG titers during longitudinal follow-up. Neutralizing antibody activity was considerably more stable than S1 and NCP IgG titers, and both reach a plateau after approximately 100 days. The sVNT proved to be not only highly specific, but also more sensitive than the specificity-focussed two-tiered serology approach. Our results demonstrate the high specificity of two-tiered serology testing and highlight the sVNT used as a valuable tool to support modelling of SARS-CoV-2 transmission dynamics, complement molecular testing and provide relevant information to individuals.

## Introduction

The novel coronavirus SARS-CoV-2 is the causative agent of the worldwide pandemic of coronavirus disease 2019 (COVID-19), which has led to millions of infections with substantial morbidity and mortality [1]. COVID-19 is characterized by a range of symptoms including cough, fever, pneumonia and a characteristic loss of smell and taste [2, 3]. The clinical manifestations of COVID-19 differ considerably and range from asymptomatic or mild self-limiting disease to severe disease and death. Next to co-morbidities predisposing to severe disease, immune hyperresponsiveness appears to be a critical factor driving COVID-19 disease severity [4, 5].

In the Netherlands, the spread of SARS-CoV-2 started in the Southern provinces, likely exacerbated by regional carnival celebrations following travel to and from Northern Italy during the school holidays [6, 7]. However, due to limited capacity at the time RT-PCR testing for SARS-CoV-2 was largely restricted to hospitalized patients with suspected COVID-19 and symptomatic individuals with moderate disease that had a recent travel history to high risk areas such as Northern Italy. Even household members of RT-PCR positive individuals were advised to self-isolate but not tested. Therefore, a large number of symptomatic individuals in The Netherlands were not tested for SARS-CoV-2 up until the start of July 2020, which left many affected individuals uncertain about whether or not their symptoms were due to COVID-19. Serological testing offers a possibility to abolish this uncertainty. In an early study, it was shown that seroconversion for anti-SARS-CoV-2 IgG and IgM occurred simultaneously or sequentially within 19 days of infection in all symptomatic COVID-19 patients analyzed [8]. The receptor-binding domain (RBD) of the spike protein of coronaviruses is a particularly interesting target for serological testing since it is the target of neutralizing antibodies [9-11], and serology tests based on the detection of these antibodies are being evaluated as indicators of protective immunity.

A crucial requirement when offering individualized serological testing, however, is a very high specificity of the test(s) selected to avoid false positives. At the early stage of the pandemic, the risk and benefit of serological tests and in particular rapid tests such as lateral flow assays (LFAs) was heavily debated [12]. Next to concerns about the performance of such rapid tests, another worry was poor registration and potential misinterpretation of the results of these tests outside controlled laboratory settings [13].

Following careful verification of a range of CE-marked serological test, Innatoss started in April 2020 to offer testing for SARS-CoV-2 antibodies in a mobile lab setting using finger prick blood. LFA results were interpreted by trained staff and applied in the context of a two-tiered testing strategy to maximize specificity. This testing strategy combined an RBD-directed LFA with strong performance characteristics [14] with a highly specific anti-SARS-CoV-2 nucleocapsid protein (NCP) IgG ELISA for confirmation in follow-up serum samples. This dual test approach is in line with the recommended common practice for serologic testing of Lyme Borreliosis, which also serves to enhance specificity [15]. Pre-screening with an LFA reduces pressure on the general health care system since it abolishes the need for venous blood collection, which is then only necessary for LFA positive individuals for confirmatory ELISA.

Here, we report the verification, performance and outcomes of this two-tiered serological testing strategy applied in n=7241 individuals from mid-April to mid-August in The Netherlands. The majority of these individuals had experienced symptoms consistent with COVID-19 at least 4 weeks prior to testing. Our results demonstrate the high specificity and feasibility of this testing approach even in times of strict anti-COVID-19 lock-down measures. We further described the kinetics of anti-SARS-CoV-2 NCP and spike protein domain 1 (S1) IgG levels as well neutralizing antibodies measured using a surrogate virus neutralization test (sVNT) [16] in a subgroup of individuals that underwent diagnostic follow-up up to 20 weeks after initial infection. These data demonstrate that neutralizing antibodies are highly stable in the time frame analyzed and that the sVNT used provides not only a very specific but also highly sensitive and scalable assay to follow-up seroconverted individuals to determine functional antibody responses to SARS-CoV-2.

## Materials and methods

### Sample collection

All blood/serum samples and data reported in this article were submitted to Innatoss for diagnostic purposes. All individuals whose data are reported in this article provided written consent to the use of pseudonymized test outcomes and of surplus serum samples for additional analyses. The Medisch Ethische Toetsingscommissie Brabant, Tilburg, The Netherlands has waived the need for IRB approval of reuse of surplus diagnostics materials (NW2020-77).

For the verification of the specificity of the serological tests in March and April 2020, up to n=224 Dutch negative control serum samples (collected before November 2019) were used. These were derived from subjects tested for different diseases at Innatoss, who had given consent to re-use the sample for quality control purposes. n=85 serum samples were tested in all tests verified. To acquire positive control samples for sensitivity verification, n=21 Dutch individuals with a past positive RT-PCR for SARS-CoV-2 as well as clinical symptoms (fever, cough) were invited to participate in the verification process and donated blood by finger prick for LFA assessment. n=11 of these individuals also donated blood by venipuncture to obtain serum samples for IgG ELISA sensitivity verification; only 6 of these were also available for IgA ELISA verification.

Whole blood samples were obtained with a finger prick in Dutch individuals with suspicion of a recently experienced SARS-CoV-2 infection. A mobile laboratory unit enabled the collection of samples in various regions of the Netherlands without violating the COVID-19 behavioural guidelines. To ensure that sufficient time had passed (>4 weeks after symptom onset) and thus that antibody responses were detectable, individuals filled in a questionnaire. This questionnaire further included the onset, end and type of symptoms they experienced prior to applying for the serological test (common cold symptoms, cough, fever, pneumonia, loss of smell or taste). Individuals were encouraged to only get tested at the mobile laboratory when they had experiencing symptoms, and were required to be symptom-free for at least 2 weeks prior to testing. Additionally, some individuals without symptoms requested testing when either a family member had experienced symptoms or when larger groups were tested for screening purposes within a company.

When individuals had a positive LFA test, they were invited to undergo a venipuncture to obtain a serum sample. This sample was used for measuring anti-SARS-CoV-2 antibodies by NCP ELISA. A second serum sample was collected from interested individuals for diagnostic follow-up. This included a group of n=97 individuals from the Dutch village of Kessel in the province of Limburg, which experienced a local SARS-CoV-2 outbreak likely in the context of a meeting with hundreds of attendees on 5 March 2020.

To verify potential cross-reactivity of the serological tests, n=23 serum samples were used including 13 samples from an internal collection of Dutch donors which were sero-positive one or more of the following pathogens: *C. Pneumoniae, M. Pneumoniae, Coxiella burnetii, Toxoplasma, Legionella*, EBV, HSV-1, CMV, Parvo virus B19). Additionally, n=10 serum samples were purchased from ProMedDx (Norton, Massachusetts, USA; n=4 human anti-mouse antibody (HAMA); n=6 rheumatoid factor (RF)). The negative control serum samples used for initial verification further included n=13 with an old and n=3 with a recent *Borrelia* infection.

### Serological tests

Whole blood obtained by finger prick was tested in two commercial CE-marked LFAs: The BIOSYNEX COVID-19 BSS using the SARS-CoV-2 receptor binding domain (RBD) as a target antigen (BIOSYNEX, Fribourg, Switzerland) and the Xiamen Boson rapid 2019-nCOV IgG/IgM Combo test using both the RBD and the SARS-CoV-2 nuclear capsid protein (NCP) (Xiamen Boson Biotech, Xiamen, China). The outcomes of the LFAs were scored positive or negative based on whether the IgG/IgM and control band were visible with the naked eye. Within the visibility, a division was made between ‘clearly visible’ and “almost invisible” to distinguish in a later phase whether the intensity of the band related to the levels of antibody detected by ELISA.

Follow-up serum samples were tested with two commercial CE-marked semi-quantitative ELISAs: The EUROIMMUN SARS-CoV-2 IgG – S1 ELISA, which uses the S1 domain and the EUROIMMUN SARS-CoV-2 IgG – NCP ELISA, modified to only contain diagnostically relevant epitopes (both from EUROIMMUN, Lübeck, Germany). Some samples were additionally tested using the *recom*Well SARS-CoV-2 IgG ELISA detecting antibodies directed against SARS-CoV-2 NCP (Mikrogen, Neuried, Germany). The EUROIMMUN SARS-CoV-2 IgA – S1 ELISA was also tested. However, during verification this test showed a specificity of only 89% and was therefore excluded from further analysis. The outcomes of the ELISAs were scored as follows according to the manufacturer’s guidelines: The EUROIMMUN ELISA results are calculated as a ratio by dividing the optical density (OD) of the sample by the OD of the calibrator (Negative <0.8; 0.8 ≤ Borderline ≤ 1.1; positive >1.1). The Mikrogen ELISA outcomes are expressed in arbitrary units calculated as (OD_sample_ / OD_cut-off control_) × 20, and test results interpreted as: Negative < 20; 20 ≤ Borderline ≤ 24; Positive > 24.

A subset of left-over serum samples was additionally tested using a CE-marked surrogate virus neutralization assay (sVNT, cPass™, GenScript, Nanjing, China) for verification and comparison purposes. The cPass™ sVNT quantifies antibodies of any isotype that interfere with the binding of the SARS-CoV-2 RBD to surface-immobilized angiotensin-converting enzyme (ACE)2. The outcome of cPass™ was expressed as the relative level of inhibition with a cut-off value of 0.2 (equivalent to 20% inhibition). The relative level of inhibition was calculated as 1-(OD_sample_ /OD_negative control_).

Finally, some individual serum samples were tested for verification and cross-reactivity using the *recom*Line SARS-CoV-2 IgG immunoblot (Mikrogen, Neuried, Germany) incorporating the SARS-CoV-2 NCP, S1 domain and RBD domain as well as the NCP of the four endemic coronaviruses (CoV) 229E, NL63, OC43 and HKU1. The immunoblot score was evaluated by scoring the intensity of the bands with the naked eye as no reaction, borderline or positive.

All of these tests were performed according to the manufacturer’s instructions. Claimed and verified characteristics of the assays are summarized in **Supporting Table S1**.

### Statistical analysis

Statistical analysis was performed using GraphPad Prism v8 (San Diego, CA, US).

## RESULTS

### Selection of tests for two-tiered SARS-CoV-2 serology assessment

To select the most appropriate tests for the two-tiered SARS-CoV-2 serology assessment, a number of CE-marked diagnostic tests including two different LFAs and four different ELISAs were verified for their specificity and sensitivity using pre-corona serum samples and samples from individuals that tested positive by RT-PCR for SARS-CoV-2, respectively (**Supporting Table S1 and Supporting Figure S1A**). Based on these data, the BIOSYNEX LFA and the EUROIMMUN NCP IgG ELISA were selected as the most appropriate tests with the highest specificity.

A panel of cross-reactivity serum samples known to be sero-positive for other pathogens, human anti-mouse antibodies (HAMA) or rheumatoid factor (RF) was tested using the combination of BIOSYNEX LFA and EUROIMMUN NCP IgG ELISA. All samples were scored negative using this two-tiered approach. The only sample that tested false-positive in the BIOSYNEX LFA with positive IgM band was the one of the six tested RF samples with the highest RF concentration (**Supporting Figure S1B**). Of note, the BIOSYNEX LFA has elsewhere also been reported to cross-react with RF [14]. Given that the false positive result in the RF serum sample by LFA was subsequently not confirmed in the NCP ELISA, the combined specificity of these two tests was 100%.

Despite the high specificity of the selected tests in negative control samples, a recurring question was whether tests would cross-react with endemic ‘common’ human coronaviruses (HCoV). Our approach had been to use a large group of sera collected in 2019 prior to the SARS-CoV-2 pandemic, and we assumed that a large proportion would be sero-positive for one or several of the HCoVs. In contrast to other diseases, control sera for common coronaviruses are not easily available. To demonstrate the presence of anti-HCoV antibodies in the collection of negative control sera, we employed an immunoblot that includes NCP antigens from the four common HCoVs as well as the SARS-CoV-2 NCP-antigen, the S1 antigen and the S1-RBD domain. This immunoblot was used to assay any negative control sample that was false positive in one of the tests verified (n=17) as well as n=9 negative control samples that were negative in all other tests. All samples tested were (borderline) sero-positive for at least one of the HCoV, with the exception of a single S1 IgA false-positive sample (**Supporting Figure S1C**).

### National and regional outcomes of the two-tiered SARS-CoV-2 serology assessment

The two-tiered testing strategy to detect seroconversion for SARS-CoV-2 was offered in a mobile lab setting for finger prick LFA testing, followed by laboratory analysis by ELISA in serum obtained from those individuals with a positive LFA. In total there were n=7241 consenting individuals who were tested by LFA between 14 April 2020 and 15 August 2020, including n=3156 males (43.6%), n=4028 females (55.6%) and n=57 unspecified individuals (0.8%). The age range of subjects was 2-95 years (median 50 years, interquartile range (IQR) 40-59 years).

Due to supply chain issues for the BIOSYNEX LFA (the first shipment was confiscated by the French authorities for priority national use), the first n=1611 individuals were tested using the Boson LFA, knowing that the 2^nd^ tier NCP IgG ELISA would filter out false positives (**Supporting Figure S1A**). Once available in mid-May 2020, the BIOSYNEX LFA was phased in and used to test another n=5630 individuals (**Supporting Table S2**). Out of n=7241 individuals that were tested by LFA, n=1481 individuals tested positive by LFA (n=478 by Boson and n=1003 by BIOSYNEX). For both LFAs, 6-7% of individuals failed to provide a follow-up serum sample for ELISA before August 15 (total n=97) (**Figure 1**). The South of the Netherlands - in particular the province of North Brabant - was the center of the initial spread of infection during the first phase of the SARS-CoV-2 pandemic, mainly linked to travel to and from Northern Italy during the school holidays in the end of February 2020, followed by regional carnival celebrations [6] (**Supporting Figure S2**). In line with this, the majority of all individuals tested was from North Brabant (4300/7241 individuals; 59,4%). When analyzing the results by area, individuals were more often positive by both LFA and NCP ELISA (35.9% of those with symptoms and 8.9% of those without symptoms) in the hardly hit village of Kessel and surroundings (situated in the municipality Peel en Maas in the province of Limburg) than in the area of Amsterdam (15.4% of those with symptoms and 1.4% of those without; **Supporting Table S2** and **Figure 1**). This is in line with the much higher incidence of notified COVID-19 cases in Peel and Maas, which is particularly striking given the much higher population density in Amsterdam compared to the rural municipality of Peel en Maas (**Supporting Figure S2C**).

**Figure 1.**
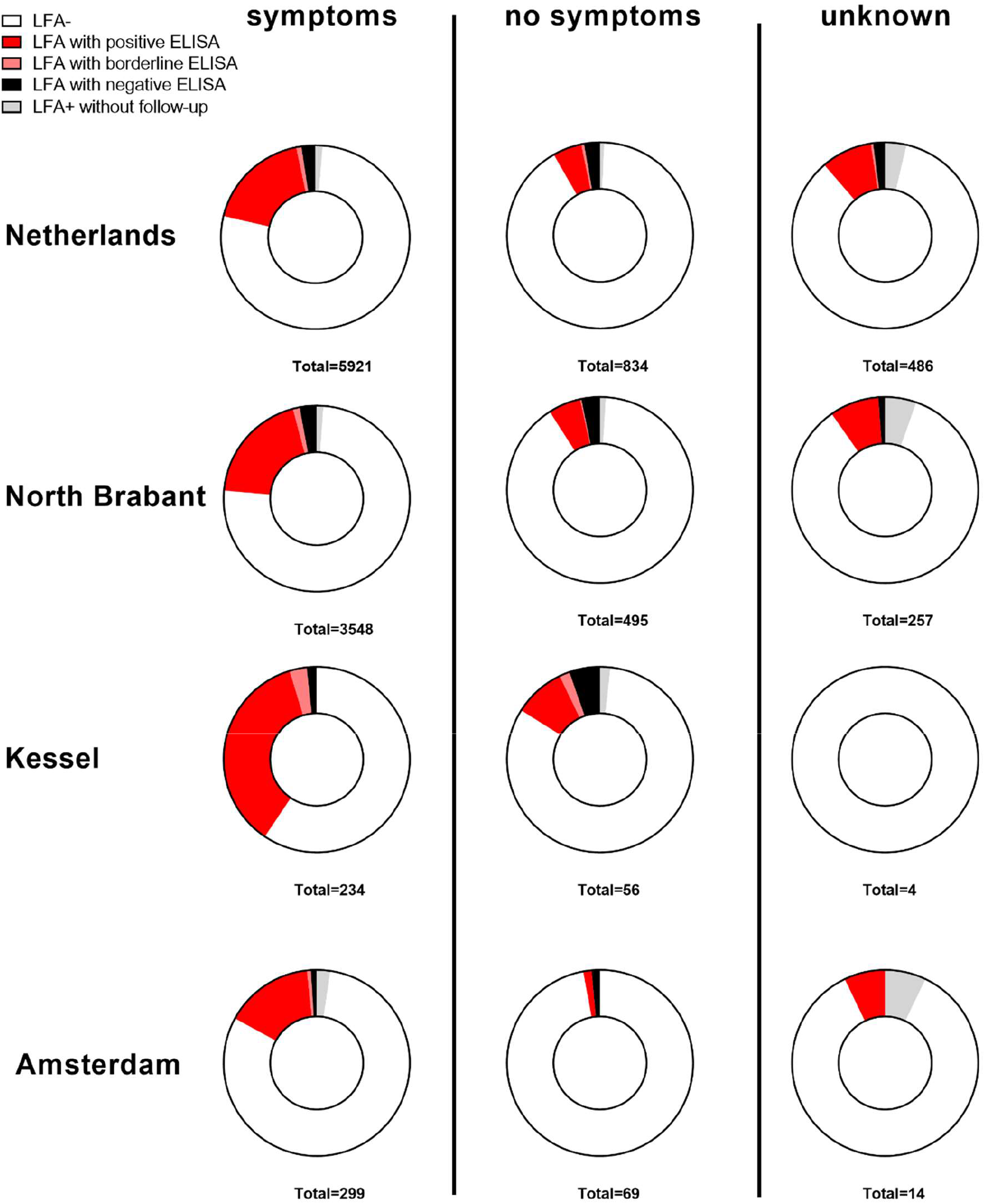
SARS-CoV-2 LFA and NCP IgG ELISA results by area. LFA and NCP ELISA results are shown for n=7241 individuals that were tested by either Boson or BIOSYNEX LFA. N=97 individuals that tested positive by LFA did not provide a follow-up serum sample for EUROIMMUN NCP IgG ELISA. Data are shown as the proportion of individuals tested in the indicated areas, stratified depending on whether or not symptoms were reported. Symptoms include common cold symptoms, cough, fever, pneumonia, loss of smell or taste.

Of note, although some individuals who requested serological testing reported symptoms as far back as October 2019, positive LFA results confirmed by ELISA were only found for individuals who reported symptoms from January 2020 onwards (**Supporting Figure S3A**). The highest proportion of individuals with self-reported symptoms that tested positive by the 2-tired LFA-ELISA strategy reported symptoms in March-May 2020 (**Supporting Figure S3B**).

Focusing on the results of the more specific BIOSYNEX LFA, a change was noted over time from samples being solitary IgM positive or IgG+IgM positive to solitary IgG positives (**Figure 2A**). LFA IgG+IgM positives could be confirmed in the EUROIMMUN NCP IgG ELISA in 94% of the cases. Confirmation of a solitary IgG positive band was lower (88.7%), except in the presence of a very clear band in the BIOSYNEX LFA (93.7%; **Figure 2B**). Moreover, the proportion of BIOSYNEX LFA positive samples that could not be confirmed by NCP IgG ELISA increased over time (**Figure 2C**).

**Figure 2.**
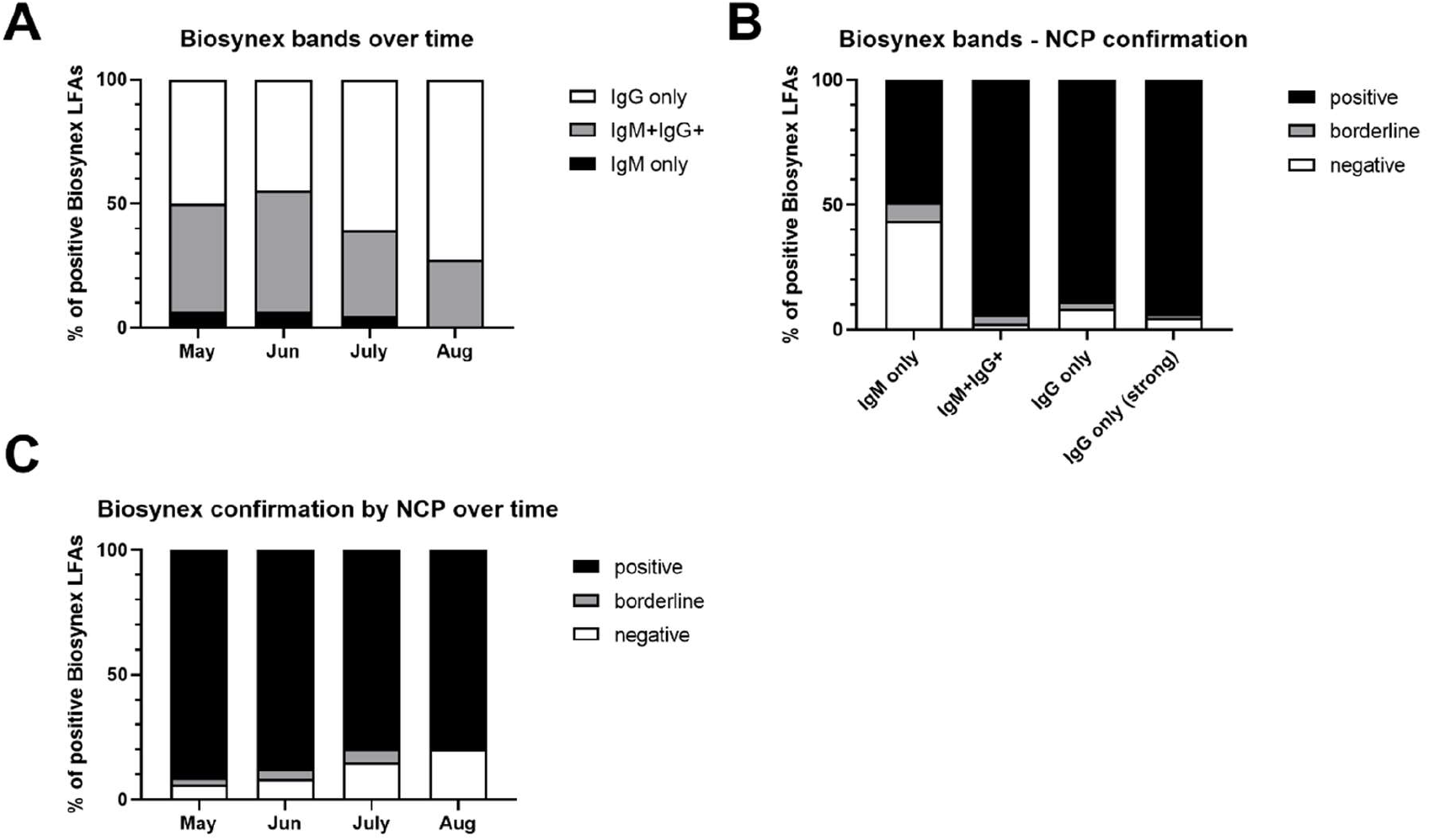
Confirmation of BIOSYNEX LFA results by EUROIMMUN NCP IgG ELISA. BIOSYNEX LFA results per month of testing (n=1003), stratified into individuals with solitary IgM or IgG bands or IgM+IgG+ bands (A). IgG levels of anti-SARS-CoV-2 NCP were determined during 2^nd^ tire serological follow-up in n=935 individuals that were positive by BIOSYNEX LFA. The proportion of samples negative, borderline or positive by NCP IgG ELISA is shown (B) for individuals with solitary IgM or IgG bands, those with both IgM+ and IgG+ bands and separately for those with a strong solitary IgG band, and for (C) all LFA IgM and/or IgG positive individuals per month of testing.

When analyzing the data regardless of whether the LFA outcome was IgM+, IgM+ and IgG+, or IgG+ and whether a Boson or BIOSYNEX LFA was used for pre-screening, then out of all LFA+, 77.9% were positive by SARS-CoV-2 NCP IgG ELISA (67,6% of Boson LFA+ and 82,8% of BIOSYNEX LFA+) and 3.9% were borderline by NCP IgG ELISA (5.2% of Boson LFA+ and 3.3% of BIOSYNEX LFA+). Moreover, 11% of positive LFA results were tested negative by NCP ELISA (21.1% of Boson LFA+ and 7.2% of BIOSYNEX LFA+; **Supporting Table S2)**. The overall lower percentages (compared to the high confirmation rates of IgG+IgM or solitary IgG+ BIOSYNEX results, **Figure 2**) are partly due to the fact that an IgG ELISA was used also to confirm results with a solitary IgM band. Moreover, these results confirm that the Boson LFA is less specific than the BIOSYNEX LFA and highlight the advantage of the two-tiered strategy of combining an LFA for screening and an ELISA for confirmation of a positive result, especially when used in combination with a less specific LFA. Not surprisingly, positive results were more common in individuals that reported symptoms (18% LFA+ and ELISA+; 0.9% LFA+ and ELISA borderline) than in those that did not (5% LFA+ and ELISA+; 0.5% LFA+ and ELISA borderline). Those who failed to report whether they had experienced symptoms or not were clearly a mixed group with 8.8% LFA+ELISA+ and 0.4% LFA+ELISA borderline.

### Stability of SARS-CoV-2 specific antibody responses

As outlined earlier, the village of Kessel and its surrounding experienced a local SARS-CoV-2 outbreak likely linked to a meeting with hundreds of participants on 5 March 2020. n=97 individuals from this area who were sero-positive in the first round of serological testing (positive LFA followed by ELISA) returned for diagnostic follow-up 19-93 days after the first measurement, enabling longitudinal analysis of antibody levels. Out of these 97 individuals, 89 reported symptoms, 7 reported no symptoms and 1 made no report. 86/89 individuals with self-reported symptoms specified a date of symptom onset, mostly in calendar weeks 9-12 (24 February – 22 March 2020; **Figure 3A**. The first serological testing was performed 39-144 days (median 65 days, IQR 55-75 days) after the onset of symptoms. There was no correlation between the time lapsed from symptom onset until the first serological test on the one hand, and the level of antibodies or antibody-mediated inhibition on the other hand (**Supporting Figure S4**). Therefore, the absolute level of antibodies and the degree of antibody-mediated inhibition did not depend on how long after the onset of symptoms the first serological test was conducted.

**Figure 3.**
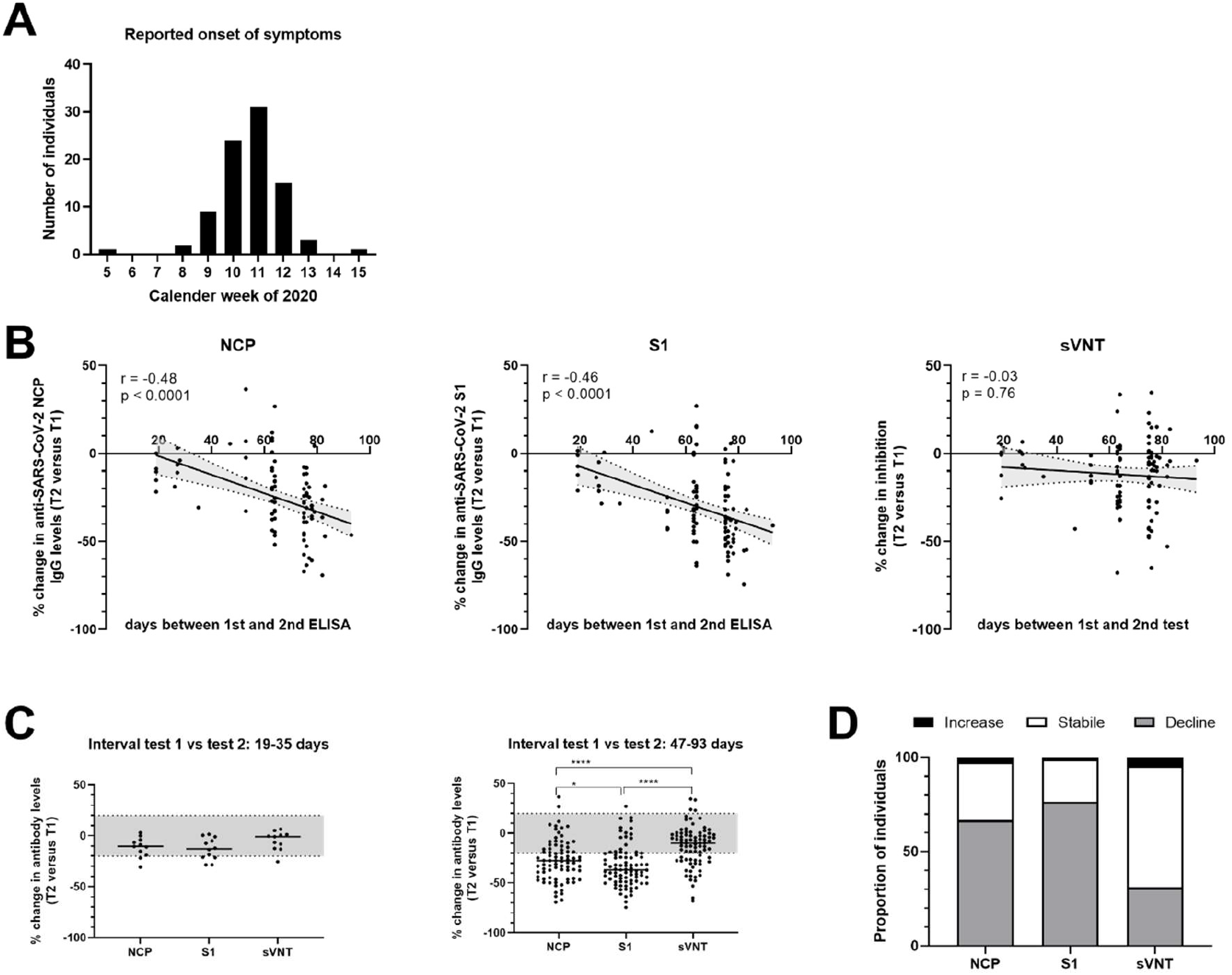
Stability of anti-NCP, anti-S1 and neutralizing antibodies. (A) The number of individuals who reported onset of symptoms is shown per calendar week. (B) The percent change in IgG levels of anti-SARS-CoV-2 NCP and S1 antibodies as well as the level of inhibition conferred by anti-SARS-CoV-2 RBD-neutralizing antibodies between the two assay time points per donor are plotted against the time between first and second ELISA/sVNT for n=97 SARS-CoV-2 sero-positive individuals regardless of presence or time of onset of reported symptoms. Data were analyzed by simple linear regression. No percent change was calculated for donors whose antibody levels at time point 1 were below the cut-off (n=5 for NCP, n=2 for sVNT). (C) Change in anti-NCP or anti-S IgG antibodies and anti-RBD neutralizing antibodies measured by sVNT in a time interval of ≤5 weeks (n=12) and >6 weeks (n=85) between first and second ELISA/sVNT. Data were analyzed by mixed-effects model analysis with Holm-Sidak’s multiple comparison test. *** p < 0.0001. (D) Proportion of individuals with declining, increasing or stable antibody levels in the group with >6 weeks (n=85) between first and second ELISA/sVNT. No percent change was calculated for those donors whose antibody levels at time point 1 were below the cut-off (n=4 for NCP, n=1 for sVNT). Changes in antibody levels were calculated by as percent change (test at time point 2 compared to test at time point 1). An increase or decrease of 20% was considered a substantial change. A change of less than +/- 20% was considered stable (grey shaded area in C).

Based on this and the fact that the outbreak occurred in a short window of time, changes in antibody levels in the interval from first to second serological assessment were analyzed for the group as a whole, regardless of the exact time of symptom onset or whether individuals reported symptoms or not. While anti-SARS-CoV-2 NCP and S1 IgG levels declined significantly over time between the first and second measurement (slope (95% confidence interval): −0.53 (−0.75 to −0.31) for NCP; −0.52 (−0,73 to −0,29) for S1; both p < 0.0001 by simple linear regression analysis), this decline was not observed for the level of antibody-mediated inhibition measured by sVNT (−0.09 (−0,33 to 0,14); p=0.43; **Figure 3B**). For further analysis, the percent change in the level of specific antibodies and antibody-mediated inhibition was calculated between the two measurement time points and an increase or decrease of 20% was considered a substantial change. When focusing on the n=85 individuals with an interval of >6 weeks (47-93 days) between the two test time points, it was evident that anti-NCP IgG and anti-S1 IgG levels declined significantly more than the antibody-mediated inhibition of RBD-ACE2 interaction. In contrast, antibody levels measured by all three testes were stable within a shorter time interval of 2-5 weeks (19-35 days) (**Figure 3C**). In this latter group of n=12 individuals, 10 reported symptoms and the first serological test was carried out much later after SARS-CoV-2 infection, 102-144 days after symptom onset (median 108; IQR 105-117 days) compared to 39-103 days after symptom onset (median 63, IQR 54-69 days) in the former group. Overall, within an interval of >6 weeks there was a decline in SARS-CoV-2 specific IgG antibodies in 65% and 75% of the individuals tested for NCP and S1, respectively. In total, 90% of individuals tested showed a decline in NCP and/or S1 IgG levels (76/85), in contrast to only 31% of the individuals when assessing antibody-mediated inhibition measured by sVNT (**Figure 3D**). The number of individuals that were scored positive at the first versus second measurement was 77/85 versus 71/85 by NCP IgG ELISA, 83/85 versus 73/85 by S1 IgG ELISA and 85/85 versus 83/85 by sVNT, stressing the stability of the sVNT results.

Already in the verification phase with negative control samples collected in 2019 as well as serum samples from SARS-CoV-2 RT-PCR+ individuals, the sVNT proved to be both highly specific and sensitive. Performance of this functional assay was further evaluated with a random selection of serum samples acquired from the mobile lab setting from n=60 LFA negative and 206 LFA positive individuals in a side-by-side comparison with the BIOSYNEX LFA, EUROIMMUN NCP and S1 IgG ELISAs (**Supporting Figure S5**). In this analysis, 8/60 LFA negative individuals had a low positive result in the sVNT assay (level of inhibition 20-54%; median 32%). It was already known from the BIOSYNEX LFA verification that this rapid test is only ∼90% sensitive, as it misses 2/20 individuals with a prior RT-PCR confirmed SARS-CoV-2 infection. In contrast, the sVNT was able to identify all 11/11 positive control samples tested. Moreover, of the 206 BIOSYNEX LFA positive serum samples, 25 were negative and 6 were borderline by NCP IgG ELISA. In contrast, only 15/206 BIOSYNEX LFA positive serum samples were negative by sVNT, all but one of which were negative by NCP IgG ELISA. These data therefore show that the highly specific and sensitive sVNT assay is able to identify additional samples that contain functional anti-SARS-CoV2 antibodies that are missed when using the BIOSYNEX RBD LFA for pre-screening or the NCP IgG ELISA for confirmation. This confirms the lower sensitivity of those two tests, alone and in combination, which were in fact selected for their excellent specificity. Not surprisingly, inhibition levels of RBD-ACE2 interaction measured by sVNT correlated more strongly with anti-S1 IgG levels than with NCP IgG level (**Supporting Figure S6**).

## Discussion

In this study, we report the verification, performance and outcomes of a two-tiered serological testing strategy combining a SARS-CoV-2 RBD-specific LFA with a confirmatory NCP IgG ELISA to assess seroconversion in n=7241 individuals, the majority of which had experienced symptoms consistent with COVID-19, but had no access to RT-PCR testing. Using this highly specific testing approach that showed no cross-reactivity with endemic HCoVs, we demonstrate that individuals reporting symptoms as far back as January 2020 showed seroconversion to SARS-CoV-2. Although individuals with past symptoms seroconverted more often than those without, a considerable proportion of 5% of asymptomatic individuals also showed seroconversion, and this number was higher in an area with a known infection cluster (8.9%) compared to a low incidence area (1.4%). Overall, 94% of individuals with a positive IgG result by BIOSYNEX LFA were also confirmed by NCP ELISA. Over time, the proportion of ELISA-confirmed LFA results declined, in line with contracting NCP IgG titers during longitudinal diagnostic follow-up in a subgroup of individuals. We further find that in contrast to S1 and NCP IgG titers, neutralizing antibody activity is a lot more stable, and that both neutralizing antibody levels and S1 and NCP IgG levels reach a plateau after approximately 100 days. The sVNT used to assess SARS-CoV-2 neutralizing antibodies proved to be not only highly specific, but also more sensitive to identify individuals with functional anti-SARS-CoV2 antibodies that are missed using the specificity-focussed 2-tiered serology assessment.

Overall, our results demonstrate the feasibility of this two-tiered testing approach to retrospectively inform individuals about the likely cause of their symptoms early during the pandemic when limited RT-PCR testing capacities left many affected individuals uncertain about whether or not their symptoms were due to COVID-19. We further demonstrate that this two-tiered strategy of combining an LFA for screening and an ELISA for confirmation of a positive result is perfectly feasible even when used in combination with a slightly less specific LFA such as the Boson Xiamen rapid test, if supplies of a more specific LFA are limited. The high specificity of the BIOSYNEX LFA makes it an interesting candidate rapid test to support molecular diagnosis or help triage suspected cases when there is shortage of RT-PCR capacities to help avoid missing true cases of COVID-19 and imposing unnecessary quarantine [17], as well as for source tracing by identifying seroconverted individuals in transmission chains that have already cleared infection and are hence not anymore RT-PCR positive.

In the meantime, the focus has shifted for individuals from the question whether or not they had been infected by SARS-CoV-2, to whether or not this prior infection and the mounted immune response would provide them with protection against future infection and thus the risk for (severe) COVID-19 and transmission of SARS-CoV-2 to others. With a potential second wave of infections coming up, this question is not only relevant on an individual level, but also for policy makers and health care authorities to determine how to optimally allocate potentially limited capacities for RT-PCR testing and more importantly both source and contact tracing. Identifying individuals with protective immunity using an easily scalable functional serological assay would allow for instance to identify individuals that do not need to undergo repeated RT-PCR testing or self-isolate when experiencing symptoms, and to release individuals from exposed clusters from quarantine measures.

Since early on in the COVID-19 pandemic, it has been suggested that serological tests could be used to issue so-called ‘immunity passports’ [18-20]. This has caused heated discussions about potential ethical, equitable and legal implications as well as public health ramifications due to potentially increased/encouraged risky behavior by ‘immunity passport’ holders [17, 21]. One often used argument in this context is that it is not yet established whether antibodies to SARS-CoV-2 confer protective immunity to further infection, what amount of antibody is needed for protection or how long any such immunity lasts. In the mean-time, a wealth of data has emerged that strongly supports both the stability (as far as it can be evaluated up to now) and protective efficacy of neutralizing antibodies to SARS-CoV-2.

In line with our findings, several studies have demonstrated that neutralizing antibodies as well as anti-RBD IgG+ memory B-cells that can produce SARS-CoV-2 neutralizing antibodies are generated and maintained in an encouragingly stable fashion for at least 3-4 months post-SARS-CoV-2 infection [22-26]. Some studies have noted a contraction in levels of anti-SARS-CoV-2 IgG antibodies including neutralizing SARS-CoV-2 antibody titers in the early convalescent phase up to 3 months after symptom onset and even complete loss in individual cases [27-29]. However, the accumulating evidence indicates that these early antibody dynamics likely only reflect the typical kinetics of a primary immune response. In particular, SARS-CoV-2-specific IgG level described for a range of individual patients show ‘text book kinetics’, with an initial peak followed by first a steep and then a much more gradual decline [28]. Overall, IgM and IgA antibody responses against SARS-CoV-2 RBD, S1 and NCP appear to contract rapidly until three months post symptom onset of COVID-19, while circulating anti-SARS-CoV-2 IgG directed against these antigens remain much more stable [23, 26, 28]. Taking into account that IgA dominates the early neutralizing antibody response to SARS-CoV-2 [30] and that neutralizing activity can be observed before an IgG response to S1 and RBD is detectable by ELISA, this strongly suggests that the initial decrease in neutralizing antibody activity simply reflects the natural contraction of the short-lived plasma blast response, while a much smaller population of long-lived plasma blasts is responsible for the continuous production of circulating IgG antibodies [23, 31, 32]. Much in line with this is our observation that RBD-specific responses detected by LFA convert from IgM to IgG over time, and that both anti-S1 and anti-NCP IgG levels decrease when re-tested up to three months after symptom onset, while the levels of antibody in follow-up samples obtained 102-144 days post symptom onset are stable. This suggests that stable antibody levels are reached within 100 days post infection. That neutralizing antibody levels showed a lesser contraction than NCP and S1 IgG levels might also be due to the affinity and avidity of anti-RBD antibodies. For very high affinity and/or avidity antibodies, a plateau of neutralizing activity may be reached, so that an initial contraction of anti-RBD antibodies is not yet translating in the same degree of decline in neutralizing capacity.

One concern is that on the longer term, antibody responses to SARS-CoV-2 might show a pattern similar to that of the four circulating (endemic) coronaviruses (HCoV-229E, OC43, NL63 and HKU1), to which most individuals are exposed to and seroconvert for the first time during childhood [33, 34]. Antibodies to HCoVs return to baseline levels within one year after natural or controlled infection [35-37] and challenge infection showed full homologous protection but only limited heterologous protection after 1 year [37, 38]. As a result, a 2-3 year cyclic re-infection pattern for endemic HCoVs is observed [34, 37, 38], which is consistent with waning neutralizing antibody titers to levels that are no longer protective. For SARS-CoV, it was observed that 50% of former patients lost detectable circulating anti-SARS-CoV antibody responses after 3 years [39]. In contrast to these total opsonizing antibodies, however, serum samples collected from recovered SARS-CoV patients 17 years after the original infection still showed neutralizing activity against anti-SARS-CoV (8/10) by sVNT [16]. This indicates that neutralizing antibody levels against this more closely to SARS-CoV-2 related coronavirus may be more long-lived. The longevity of the neutralizing antibody response to SARS-CoV-2 will need to be determined by carefully designed longitudinal follow-up studies in cohorts of seroconverted individuals like ours, ideally with prospective analysis of potential re-infections. A modelling study of SARS-CoV-2 transmission has demonstrated how the duration of immunity to SARS-CoV-2 infection determines whether or not SARS-CoV-2 enters into regular circulation after the initial pandemic wave [40]. Of note, even a duration of ‘only’ two years compared to one year of protection would already positively affect the total incidence of COVID-19 over the next five years. On a population level, sero-epidemiological data on neutralizing antibody levels and their longevity are therefore crucial to support modelling the impact of preventative measures and post-pandemic transmission dynamics [40, 41].

In regards to protective efficacy, several animal models have provided evidence that SARS-CoV-2 infection induces protective immunity against re-challenge in rhesus macaques, Syrian hamsters and ferrets [42-45], and protection from disease could be linked to neutralizing antibodies (either naturally induced or passively transferred) in the Syrian golden hamster model [44, 46, 47]. In humans, neutralizing antibodies were shown to correlate with protection from SARS-CoV-2 during a high attack rate fishery vessel outbreak, in which three sero-positive crewmembers were amongst the few that did not contract infection [48]. And even in the event of re-infection, symptoms are likely to be milder or even absent, as reported for the first documented case for SARS-CoV-2 reinfection [49]. Consequently, passive immunization using convalescent plasma or monoclonal antibodies for SARS-CoV and SARS-CoV-2 is also pursued as therapeutic approach to treat critically ill COVID-19 patients [50-57] and antibody neutralization assays are used as a key read-out in SARS-CoV-2 vaccination trials in non-human primates and human clinical trials [58-62].

Finally, in hospitalized COVID-19 patients, it was further observed that upon seroconversion, shedding of infectious SARS-CoV-2 dropped rapidly to undetectable levels. Infectious virus could not be isolated from respiratory tract samples once patients had a serum neutralizing antibody titer of at least 1:80 measured by plaque reduction neutralization test (PRNT), which may be due to the fact that infectious virions are still produced but directly neutralized by antibodies in the respiratory tract. Whether the same holds true for individuals with mild disease remains to be determined. Nevertheless, the authors suggested that both quantitative viral RNA load assays and serological assays should be used to monitor individuals to discontinue or de-escalate infection prevention and control precautions [63].

While PRNTs are the gold standard for assessing functional antibody activity, these assays are labor-intensive and require a biosafety level 3 laboratory. In contrast, the sVNT assay employed here is scalable to high throughput use and has shown good agreement with PRNT [16], and has hence been highlighted as an interesting alternative to PRNT to quantify functional antibodies [64]. This sVNT also used in a recently published Australian study evaluating a range of serological tests for SARS-CoV-2 [65]. This study reported slightly lower sensitivity values for the sVNT, likely due to the fact that serum samples from convalescent Australian COVID-19 patients were collected at an earlier time point in the convalescent phase. Indeed, they report a considerable increase in sVNT sensitivity depending on whether serum samples were collected within 14 days or after 14 days of symptom onset. In our study, all but 2/235 of the individuals with a reported date of symptom onset that were tested by sVNT provided their serum samples at least 4 weeks after symptom onset (median 62 days, IQR 51-86 days). Ultimately, a key question to be answered in future studies is how the level of inhibition measured by sVNT at a given serum dilution compares to the level of neutralizing antibody required at which shedding of infectious virus is reduced to undetectable levels. This information will be highly valuable for the interpretation of the sVNT test outcome in regards to protection from future infection in an ‘immunity passport’ scenario. Given that it is not yet known how long neutralizing antibodies to SARS-CoV-2 are stable and that this may also differ on an individual level, periodic re-testing will likely be required.

There are some limitations to our study. Firstly, it should be highlighted that our data do not reflect the seroconversion dynamics in the general population, since we encouraged individuals only to get tested if they had experienced symptoms consistent with COVID-19. Even amongst this group, the proportions of seroconversions we find are not representative for the whole of the Netherlands: Although tests were performed in almost every region of the Netherlands, the majority of tests were carried out in North Brabant (the most heavily affected area in March/April 2020) and Amsterdam. Secondly, using the specificity-focussed two-tiered serological testing approach applied here, we were bound to miss some individuals that had seroconverted. This is exacerbated over time by the fact that NCP IgG levels contract and thus an increasing proportion of individuals that show a positive result by RBD-LFA cannot be confirmed by NCP IgG ELISA. Using the sVNT in combination with the RBD-LFA for pre-screening may partially alleviate this limitation. However, seeing the high scalability of the sVNT in combination with both its high specificity and sensitivity (provided samples are collected long enough after symptom onset), it can also simply be used as a stand-alone screening tool. Thirdly, as any serological screening approach, the small proportion of individuals which fail to seroconvert [22, 66] and instead only mount a cellular immune response to SARS-CoV-2 [67] will logically not be identified. Given that the degree of the antibodies seems to be related also to disease severity, this may be particularly true for individuals with mild or asymptomatic infection [27, 68]. Finally, while our data indicate that stable antibody levels are reached 100 days post infection, the group of individuals that was assessed >100 days post symptom onset consisted of a mere n=12 individuals. Therefore, analysis in larger groups is needed to support this finding.

In conclusion, we herein present a careful analysis of the performance of a two-tiered serological testing approach to assess seroconversion and thus retrospectively confirm exposure to SARS-CoV-2. Our data support that the highly specific LFA used in this combination approach may be useful also in other settings, for instance to support or complement molecular testing and source tracing of SARS-CoV-2 infection in transmission chains. Finally, we confirm other studies showing the stability of neutralizing antibodies to SARS-CoV-2, and provide the first longitudinal assessment of these antibodies using a highly scalable sVNT. This high specificity and sensitivity assay could thus be a valuable tool for serological follow-up in large cohorts to support modelling of future SARS-CoV-2 transmission dynamics, support health authorities and provide relevant serological information to individuals.

## Data Availability

The data that support the findings of this study are available from the corresponding author upon reasonable request

## Acknowledgements

We acknowledge the continuous support of Stichting IMtest and in particular Dorien van Doorn on the logistic challenges that we faced; Luc van Galen for setting up the MoLabs; Sanne van’t Hullenaar and Saskia Theunisse for coordinating the MoLab appointments; Guido van de Wiel for setting up the MoLab IT infrastructure; Kim Plaum for blood collections in the verification phase; and the MoLab team consisting of Anna van Mölken, Laura van den Ende, Sarah van den As, Willem Baijens, Laurens Bierens, Laurie Tiehuis, Koen van Maren, Sjoerd van der Burg, Sander Colijn, Daan van den Nieuwenhof and Luc van Galen for running more than 8000 lateral flow tests and helping with analyzing the clinical data. Jasper van Galen was instrumental in realizing timely reporting to all the individuals involved. Finally, we acknowledge Liza van Oort (Mediphos Medical Supplies) for support in a challenging environment.

## Authors contributions

Conceptualization and design: AG, MLCEK. Acquisition of data: DWAvdN, APFS, ESD, LSvG. Analysis and interpretation of data: AG, AS, DWAvdN, MLCEK. Drafting the article: AS, DWAvdN. Revision of article: AG, MLCEK. All authors have read and approved the final manuscript.

