## Supporting information for "Two-tiered SARS-CoV-2 seroconversion screening in the Netherlands and stability of nucleocapsid, spike protein domain 1 and neutralizing antibodies"

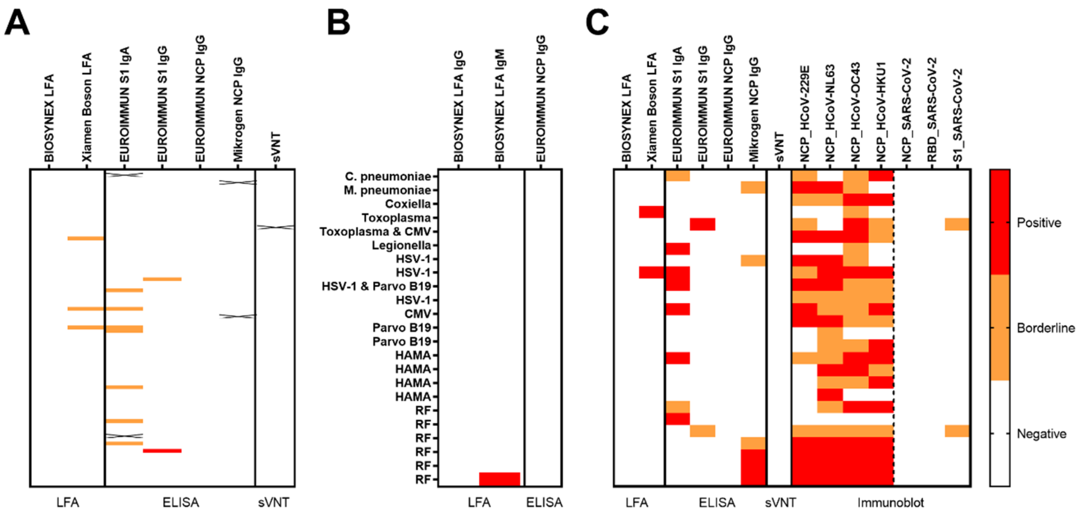


***Figure S1 – SARS-CoV-2 LFA, ELISA and immunoblot results for selected negative control and cross-reactivity samples.*** *(A) n=85 negative control serum samples collected in 2019 were assayed in two LFAs, 4 ELISAs and the sVNT. (B) n=23 cross-reactivity serum samples were tested by BIOSYNEX LFA and EUROIMMUN NCP IgG ELISA, followed up by immunoblot for the single BIOSYNEX positive sample. (C) An immunoblot was used to assay n=17 negative control sample that were false positive in one of the tests verified as well as n=9 negative control samples that were negative in all other tests. Each row shows the test outcome for one donor. Crosses indicate tests that were not performed for a given sample.*

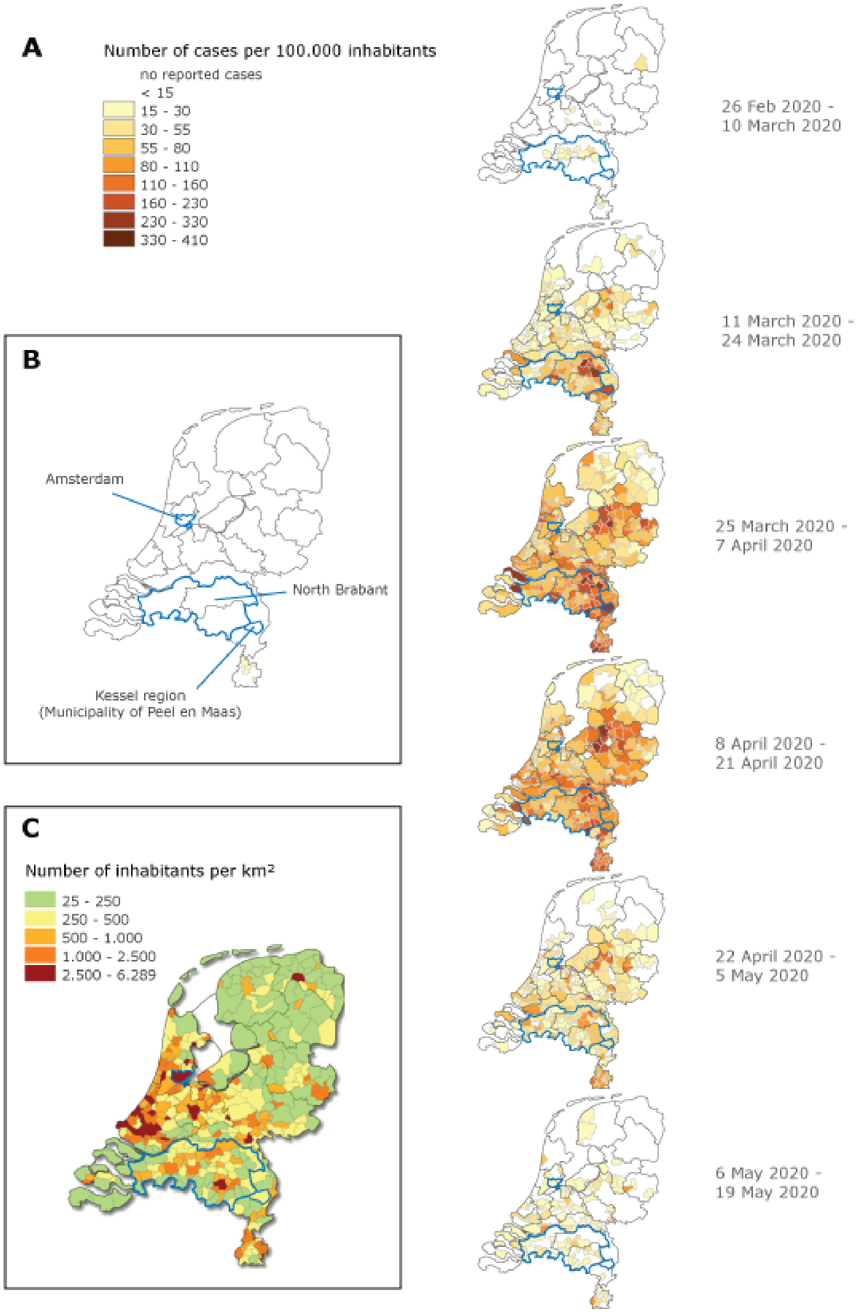


***Figure S2 – Incidence of reported COVID-19 cases and population density in the Netherlands.*** *(A) The number of reported COVID-19 cases per municipality in the Netherlands over time in the period of the first outbreak, shown per 100,000 inhabitants. Data are based on SARS-CoV-2RT-PCR positive swaps obtained predominantly from hospitalized individuals or symptomatic individuals with a recent travel history to high risk areas. (B) The province of North Brabant and the municipalities of Peel en Maas and Amsterdam are highlighted by blue outlines. (C) Population density in the different municipalities of the Netherlands in 2015. All data are derived from websites by the Dutch Institute for Public Health and Environment (https://www.rivm.nl/coronavirus-covid-19/actueel and https://www.volksgezondheidenzorg.info).*

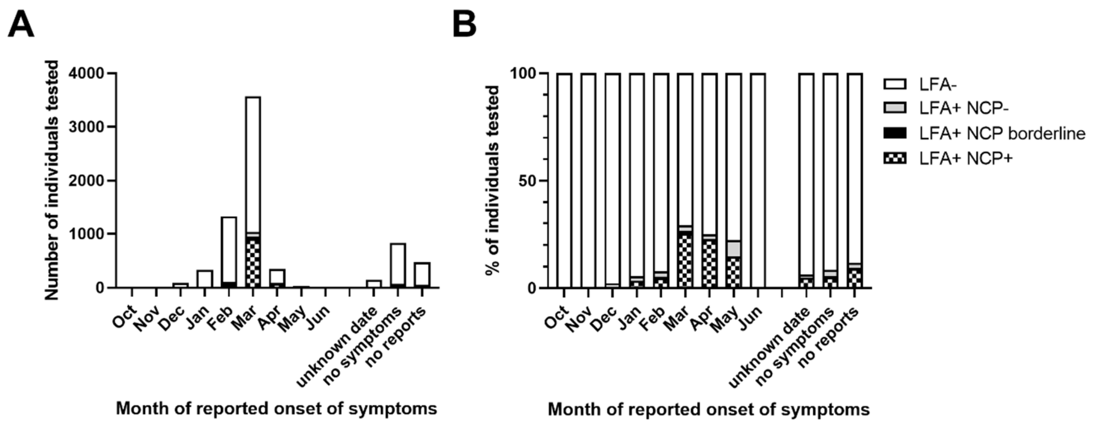


***Figure S3 – SARS-CoV-2 LFA and NCP IgG ELISA results depending on month of symptom onset.*** *LFA and NCP ELISA results are shown for n=7144 individuals that were tested by either Boson or BIOSYNEX LFA and completed follow-up by EUROIMMUN NCP ELISA if tested positive by LFA. n=97 individuals did not provide a follow-up serum sample for ELISA in time and are not included into this analysis. Data are shown as the number (A) or proportion (B) of individuals negative or positive by LFA that had a negative, borderline or positive result in the NCP IgG ELISA, stratified by month of reported symptom onset. Out of the n=7144 individuals, a number of individual reported symptoms without a specific date of onset (n=144), no symptoms (n=827) or made no report about whether or not they had symptoms (n=470).*

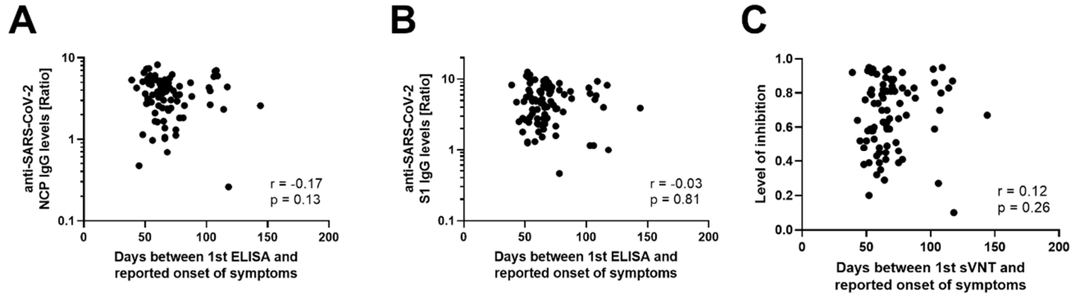


***Figure S4 – Antibody levels relative to time of symptom onset.*** *IgG levels of anti-SARS-CoV-2 NCP (A) and S1 (B) antibodies as well as the level of inhibition conferred by anti-SARS-CoV-2 RBD-neutralizing antibodies (C) are plotted against the time between first ELISA/sVNT and reported date of symptom onset for n=86 individuals. Data were analyzed by Spearman correlation.*

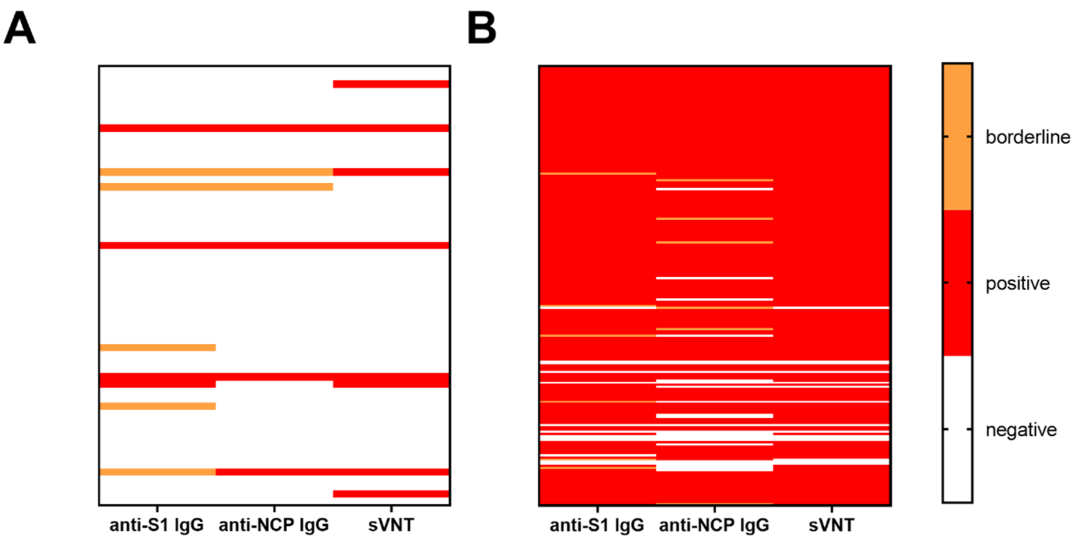


***Figure S5 – SARS-CoV-2 S1 or NCP specific and neutralizing antibodies in BIOSYNEX LFA positive and negative individuals.*** *A selection of n=266 serum samples from individuals tested in the mobile lab was assessed side-by-side by BIOSYNEX LFA, EUROIMMUN S1 and NCP IgG ELISA and sVNT. ELISA and sVNT results are shown for (A) n=60 LFA negative and (B) n=206 LFA positive serum samples.*

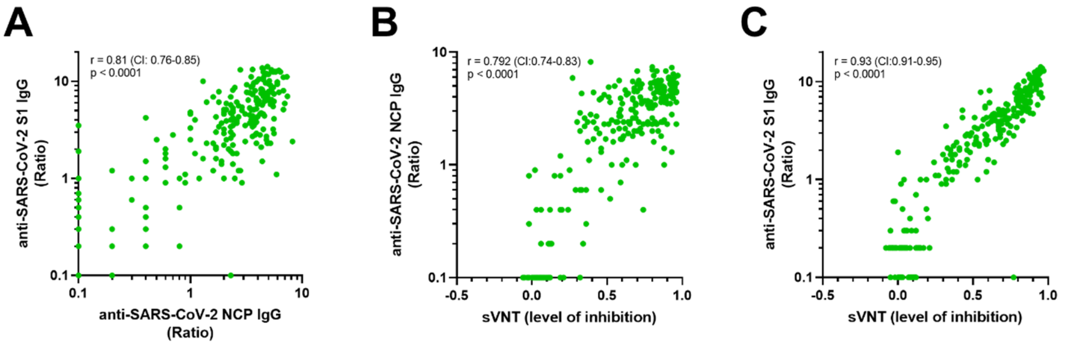


***Figure S6 – Correlation of SARS-CoV-2 S1 or NCP specific and neutralizing antibodies.*** *A selection of n=266 serum samples from individuals tested in the mobile lab was assessed side-by-side by EUROIMMUN S1 and NCP IgG ELISA and sVNT. Spearman correlation analysis is shown for (A) anti-S1 vs anti-NCP IgG levels, (B) anti-NCP IgG levels versus inhibition by sVNT and (C) anti-S1 IgG levels versus inhibition by sVNT.*

**Tables**

***Table S1 - Claimed and verified characteristics of serological tests***

| | **Claimed by manufacturer** | | **Verified** | |
|---|---|---|---|---|
| | Sensitivity | Specificity | Sensitivity[1] | Specificity[2] |
| **LFA** | | | | |
| Xiamen Boson | IgG and/or IgM: 87.8% (65/74) | IgM: 99.7% (304/305) IgG: 99.3% (303/305) | IgG and/or IgM: 90.5% (19/21) | IgM: 99% (99/100) IgG: 98% (98/100) |
| BIOSYNEX | IgM: 91.8% (74/81) IgG:100% (77/77) | IgM: 99.2% (372/375) IgG: 99.5% (367/369) | IgG and/or IgM: 90.0% (18/20) | IgM: 100% (223/224) IgG: 100% (224/224) |
| **ELISA** | | | | |
| EUROIMMUN IgA – S1 | 100% (5/5)[1] | 92.5% (185/200) | 83% (5/6) | 89% (76/85) |
| EUROIMMUN IgG – S1 | 90.0% (27/30) | 100% (80/80) | 100% (11/11) | 98% (87/89) |
| EUROIMMUN IgG – NCP | 86.7% (26/30) | 99.8% (1036/1038) | 100% (11/11) | 100% (115/115) |
| Mikrogen recomWell IgG – NCP | 100% (28/28) | 98.7% (296/300) | 100% (11/11) | 97% (179/185) |
| **Immunoblot** | | | | |
| Mikrogen recomLine IgG | 96.3% (52/54) | 98.8% (564/571) | 100% (2/2) | 100% for SARS-CoV-2 NCP and RBD (26/26). 92% for S1 (24/26) |
| **Surrogate Virus Neutralization Test** | | | | |
| Genscript SARS-CoV-2 sVNT (cPass™) | 93.3% (56/60) | 100% (97/97) | 100% (11/11) | 100% (84/84) |

1. These percentages have a large confidence interval due to the small number of samples available. The focus in this study was to prevent false positive results, which made sensitivity of lesser importance.

2. In all ELISA results for specificity calculations, a borderline outcome was interpreted as a positive outcome.

*Table S2 – SARS-CoV-2 LFA and NCP IgG ELISA results by area*

| | **The Netherlands** | | | **North Brabant[5]** | | | **Kessel[5]** | | | **Amsterdam[5]** | | |
|---|---|---|---|---|---|---|---|---|---|---|---|---|
| | BIOSYNEX | Xiamen Boson | **Total** | BIOSYNEX | Xiamen Boson | **Total** | BIOSYNEX | Xiamen Boson | **Total** | BIOSYNEX | Xiamen Boson | **Total** |
| Total tested | 5630 | 1611 | **7241** | 2895 | 1405 | **4300** | 84 | 210 | **294** | 323 | 59 | **382** |
| **Symptoms[1]** | | | | | | | | | | | | |
| Total | 4654 | 1267 | **5921** | 2394 | 1154 | **3548** | 70 | 164 | **234** | 268 | 31 | **299** |
| LFA positive | 907 | 426 | **1333** | 482 | 396 | **878** | 16 | 79 | **95** | 46 | 12 | **58** |
| [no follow-up[3]] | [48] | [25] | **[73]** | [21] | [20] | **[41]** | [0] | [0] | **[0]** | [5] | [2] | **[7]** |
| ELISA positive | 771 | 297 | **1068** | 417 | 274 | **691** | 15 | 69 | **84** | 37 | 9 | **46** |
| [borderline[4]] | [27] | [25] | **[52]** | [16] | [25] | **[41]** | [1] | [6] | **[7]** | [2] | [0] | **[2]** |
| **No symptoms** | | | | | | | | | | | | |
| Total | 526 | 308 | **834** | 273 | 222 | **495** | 13 | 43 | **56** | 41 | 28 | **69** |
| LFA positive | 32 | 43 | **75** | 15 | 35 | **50** | 3 | 7 | **10** | 1 | 1 | **2** |
| [no follow-up] | [3] | [3] | **[6]** | [2] | [3] | **[5]** | [0] | [1] | **[1]** | [0] | [0] | **[0]** |
| ELISA positive | 22 | 20 | **42** | 10 | 18 | **28** | 2 | 3 | **5** | 1 | 0 | **1** |
| [borderline] | [2] | [0] | **[2]** | [1] | [0] | **[1]** | [1] | [0] | **[1]** | [0] | [0] | **[0]** |
| **Unknown[2]** | | | | | | | | | | | | |
| Total | 450 | 36 | **486** | 228 | 29 | **257** | 1 | 3 | **4** | 14 | 0 | **14** |
| LFA positive | 386 | 9 | **395** | 31 | 8 | **39** | 0 | 0 | **0** | 2 | 0 | **2** |
| [no follow-up] | [17] | [1] | **[18]** | [13] | [1] | **[14]** | [0] | [0] | **[0]** | [1] | [0] | **[0]** |
| ELISA positive | 37 | 6 | **43** | 17 | 5 | **22** | 0 | 0 | **0** | 1 | 0 | **1** |
| [borderline] | [2] | [0] | **[2]** | [0] | [0] | **[0]** | [0] | [0] | **[0]** | [0] | [0] | **[0]** |

1. Symptoms reported include common cold symptoms, cough, fever, pneumonia, loss of smell or taste

2. No answers were provided in the questionnaire about the presence or absence of symptoms

3. Number of all individuals that tested positive by LFA [number of individuals within that group which not provide a serum sample for follow-up ELISA]

4. Number of LFA+ individuals with a positive outcome by follow-up NCP IgG ELISA [additional number of LFA+ individuals with a borderline outcome by follow-up NCP IgG ELISA]

5. North Brabant: Postcode starting with 4 or 5; Kessel: Postcode starting with 599 or 598; Amsterdam: Postcode starting with 10 or 11.
